# HIV risk score and prediction model in the United States: A Scoping Review Protocol

**DOI:** 10.1101/2023.12.18.23300171

**Authors:** Maitri Patel, Krishna Vaddiparti, Parisi Christina, Aislin Woody, Robert Cook, Mattia Prosperi, Courtney Pyche, Christa Cook, Yiyang Liu

**Author notes:** **Corresponding author:** Yiyang Liu, MPH, PhD, 2004 Mowry Road, Room, Research Assistant Professor, Department of Epidemiology, University of Florida, PO Box 100231, Gainesville, FL 32610., |.

## Abstract

Human immunodeficiency virus (HIV) remains a significant public health concern in the United States, affecting approximately 1.2 million individuals, with a substantial portion unaware of their infection status. Identifying individuals at elevated risk of HIV infection through predictive models holds promise for enhancing public health efforts. A preliminary search from PubMed revealed a handful of studies focused on developing HIV risk prediction models or risk scores, each employing varying methodologies such as logistic regression and machine learning. These studies targeted a diverse population including men who have sex with men, emergency department visitors, and the general population, drawing data from surveys, surveillance, and electronic health records. Despite these individual efforts, there is a notable absence of comprehensive review papers summarizing the outcomes of these studies. Addressing this gap, this scoping review systematically identifies and synthesizes results from existing predictive models for HIV risk. The primary objective is to determine the variables used in HIV risk scoring and prediction models, contributing to a more comprehensive understanding of HIV risk assessment. This protocol describes the thorough procedure for conducting a scoping review. It outlines the inclusion and exclusion criteria for relevant studies and offers the search methods that will be used in PubMed, EMBASE, and CINAHL. Detailed paper screening, data extraction, and risk of bias assessment process were described.

## Introduction

Human immunodeficiency virus (HIV) affects around 1.2 million persons in the US, with 14% of those individuals uninformed of their infection status. While addressing the detection and treatment of HIV-infected people continues to be a major public health priority, limiting the transmission of the virus is equally crucial (Chou et al., 2019; Delaney & DiNenno, 2021). HIV pre-exposure prophylaxis (PrEP), a preventive approach involving the use of antiretroviral medications by high-risk individuals, has emerged as a key strategy in reducing new HIV infections. Moreover, routine HIV testing among the high-risk population would contribute to early detection of the infection, facilitate timely access to treatment and care, and contribute to reduced HIV transmission within the community.

A disease risk predictive model is a statistical or computational tool used to estimate and forecast the occurrence of a specific disease (e.g., new HIV diagnosis) within a population over a defined period. These predictive models can generate a disease risk score, which represents a quantitative measure of an individual’s probability of acquiring the health condition. Using the predictive models and HIV risk score to identify individuals at elevated risk of HIV infection holds significant promise for improving HIV prevention and testing efforts. The automated identification of individuals at elevated risk has the potential to enhance public health efforts and curb HIV transmission. (Baeten et al., 2012; Krakower et al., 2019; Marcus et al., n.d.). To optimize the public health advantages of PrEP and routine HIV testing, it is crucial to make sure that it is available to people who are most at risk for contracting HIV.

A preliminary search on PubMed reveals papers on developing, validating, or updating predictive models or risk scores to identify individuals at higher HIV risk in the US. A diverse patchwork of studies spanning a wide range of approaches and populations under evaluation has formed in the field of HIV risk prediction models. However, great heterogeneity was observed in these studies. The studies conducted by Muttai et al., Goetz et al., Burns et al., and Tordoff et al., showcased the global scope of research efforts, spanning from Western Kenya to the Southern United States and focusing on specific demographics like men who have sex with men. Duthe et al., emphasized the international applicability of predictive models, utilizing machine learning and Electronic Health Record (EHR) data to efficiently identify patients at risk of HIV infection. Marcus et al., expanded on predictive modeling, developing a comprehensive HIV risk assessment tool for men who have sex with men, incorporating demographic and behavioral factors, underscoring the necessity for inclusive risk assessment strategies. Ridgway et al., pioneered the implementation of electronic risk scores in Emergency Departments, showcasing the practical application of automated risk assessments and linking high-risk individuals to timely preventive care. This amalgamation of diverse methodologies, targeted populations, data resources, and risk assessment strategies highlights the complexity of HIV risk prediction and prevention efforts.

Therefore, a comprehensive scoping review becomes imperative to synthesize these varied approaches, exploring the predictors included in existing HIV risk prediction models and risk scores in the US. By addressing primary research questions on the predictors, overall model performance, and the incorporation of social determinants of health and community-level factors, this scoping review aims to provide an in-depth understanding of the existing landscape and pave the way for future advancements in HIV risk prediction and prevention.

We aim to achieve the following objectives through this scoping review:

Primary: What are the predictors included in existing HIV risk prediction models and risk scores in the US?

Secondary: 1) How do these predictive models perform overall, and do they show variations by racial and gender differences? 2) To what extent do these models incorporate social determinants of health and community-level factors in their design and assessment?

### Eligibility criteria

#### Population

The target population of this scoping review is adolescents and adults (aged thirteen or older) at risk of HIV of all genders, sexes, and ethnicities at risk of HIV in the United States (US).

- We will exclude studies that only include participants outside of the US.
- We will only include studies that have the HIV outcomes (defined below under Concept) measured among adolescents and adults. We will exclude studies that only measure HIV outcomes among people below the age of 13, despite including people aged 13 or older in the study sample (e.g., predicting HIV risk among infants even though adult patients were included in the study sample).
- Studies with participants who were already diagnosed with HIV at baseline will be excluded.

#### Concept

- We will include studies focusing on diagnostic prediction models that aim to develop, validate, or update a multivariable prediction model with the outcome being HIV incidence, new HIV diagnosis, new HIV infections, or first positive HIV testing.
- We will exclude studies that involve the development or validation of risk scores of comorbid conditions in the HIV population. (e.g., risk score of cardiovascular disease among People With HIV).
- We will exclude risk factor studies that aim to identify risk or protective factors independently associated with or having causal effects on HIV risk will be excluded.
- We will exclude Prediction model impact studies, which evaluate the effect of using a predictive model to guide patient care compared to not using such a model.
- We will exclude computable phenotype studies to identify a cohort of patients newly diagnosed with HIV.

#### Context

- We will include full-text peer-reviewed published original studies.
- Papers published from the year 2010 to 2023 will be included.
- We will exclude abstract-only publications, review articles, or publications from non-peer-reviewed sources.
- We will exclude manuscripts that lack abstracts for title and abstract screening or full text for full-text screening (after using the interlibrary loan system offered by our university).
- We will exclude studies not published in English.

### Search Strategy

The literature search was conducted in three electronic databases: PubMed, EMBASE and CINHAL.

The search approach will involve keyword groupings that are specific to each database about two concepts: 1. HIV/PrEP (search terms 1 and 2) and 2. Prediction Model or Risk Score (search term 3).

Details of the search terms are included in the table. This list will be used as a starting point for study selection and will be revised during the full-text screening process.

**Table 1:**
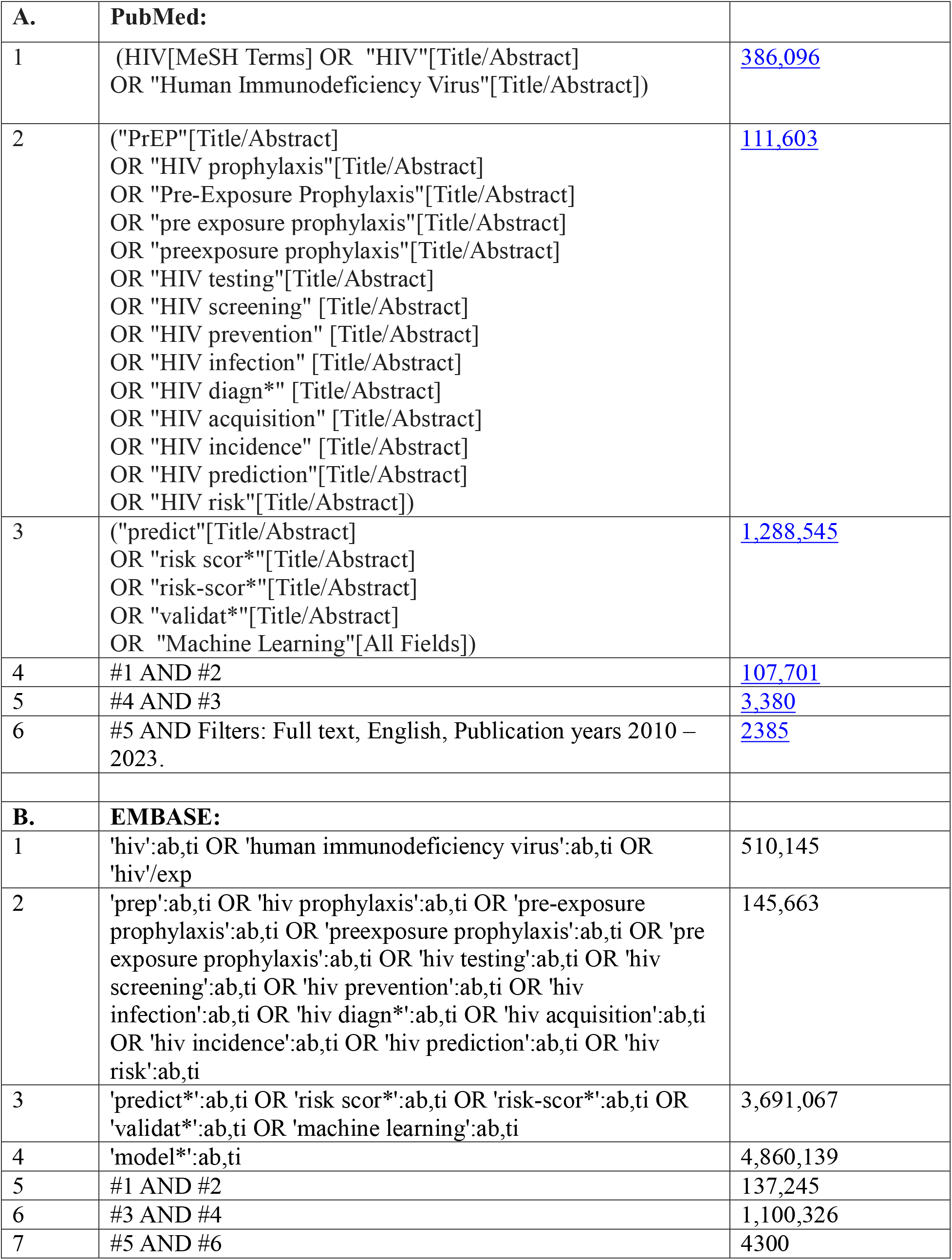
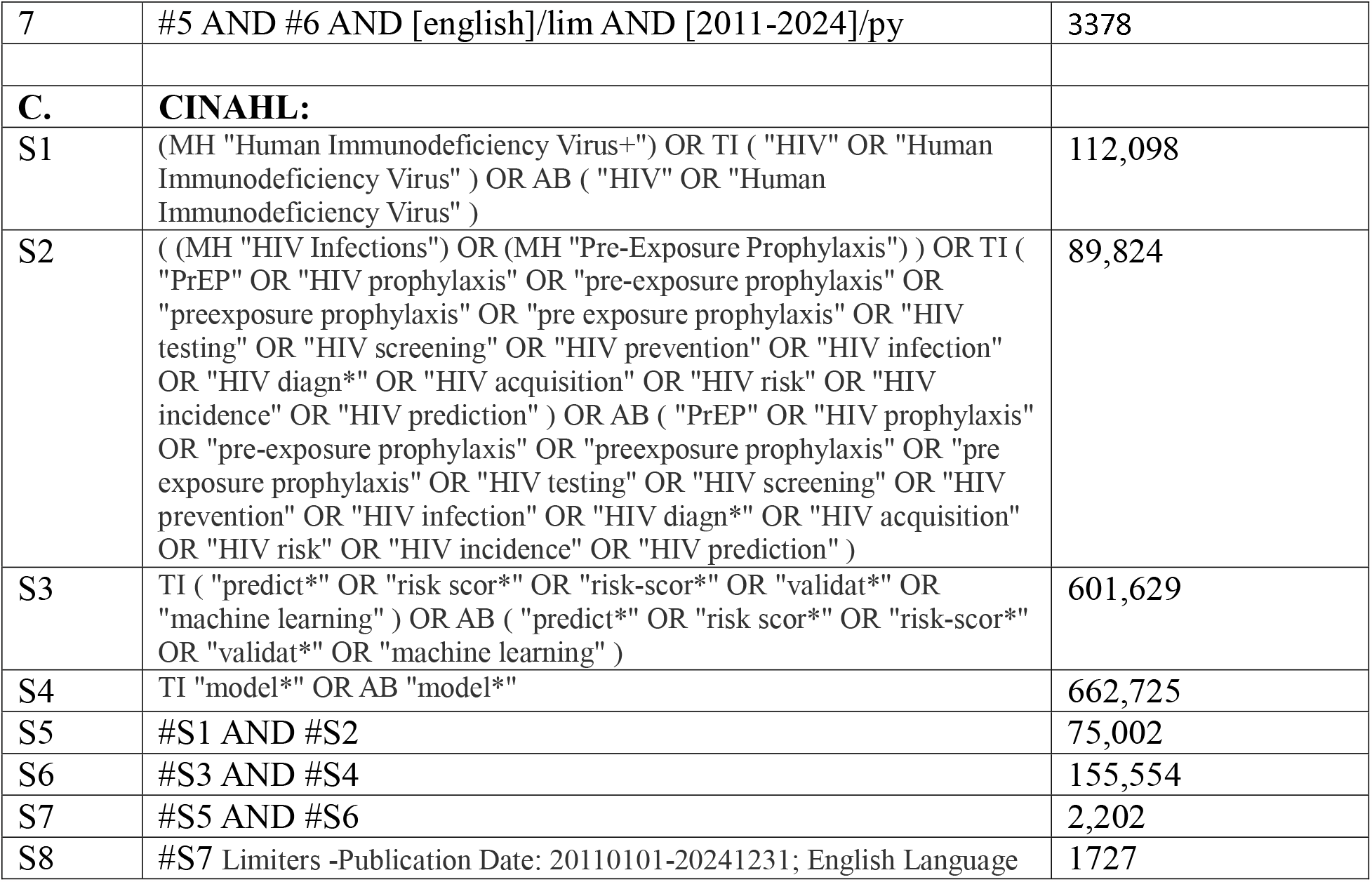
A – PubMed search terms and search results, B – EMBASE search terms and search results, C-CINAHL search terms and search results.

### Selection Process

We will export all identified papers through the search strategy into a systematic review tool, such as Covidence and Rayyan, to remove any duplications. The first stage of selection entails abstracts and title screening. Two reviewers will independently assess all titles and abstracts for inclusion. Any discrepancies that arise will be discussed between the reviewers to reach a consensus. For unresolved discrepancies, a third senior reviewer/Principal Investigator (PI) will be consulted. Additionally, a third senior reviewer will screen the first 5-10% of abstracts/titles alongside the two primary reviewers to assess concordance and screening quality.

The second stage of the selection process involves full-text screening. This process is similar, but this time we will assign reasons for exclusion for each excluded study. Furthermore, we will identify potential inclusions by reviewing the citations of already included papers. Subsequently, a flowchart illustrating the paper identification process will be constructed.

### Data extraction process

Guided by the Checklist for critical Appraisal and data extraction for systematic Reviews of prediction Modelling Studies (CHARMS), we will extract the following information from included papers: source of data (survey data, surveillance data, registry data, EHR, Claims), study design (e.g., cohort study, case-control, case-cohort, cross-sectional), participant descriptions (recruitment location, sample size (including number of total participant, number of outcomes/event, events per variable), and basic demographics), definition of outcome (including time of outcome occurrence and duration of follow-up period), number and type of predictors (demographics, behavioral factors, clinical characteristics, social determinants of health, community-level factors and other factors), candidate predictor measurement (including timing of measurement), handling of missing data (number of people with missing value for predictors and outcome, complete-case analysis or imputation), modeling methods (e.g., logistic regression, survival analysis, or machine learning techniques), final multivariable models (e.g, basic, extended, simplified models or model for any specific subgroups; including predictor weights, regression coefficients, and risk score, and model performance (overall and by sex or race if applicable; e.g., C-statistics, sensitivity, specificity, positive and negative predictive values; cutoff-point used).

Before beginning data extraction, a pilot test will be conducted using a small subset of studies (e.g., 2 papers) to ensure consistency and agreement among team members. Any discrepancies will be resolved through discussion and refinement of the extraction forms. Team members will independently extract data from each included study using the standardized data extraction forms. Any discrepancies or disagreements in data extraction will be resolved through discussion and consensus among team members. In cases where consensus cannot be reached, a third senior reviewer/PI will be consulted. Regular meetings will be scheduled to review and verify data extraction procedures, ensuring accuracy and completeness.

### Risk of bias assessment

Two reviewers will independently carry out quality and risk of bias assessment. We will use PROBAST (Prediction model Risk Of Bias ASsessment Tool) to assess the risk of bias and any concerns regarding the applicability of diagnostic model studies. (PROBAST) We will utilize PROBAST to assess studies of model development and model validation. Two reviewers will send and discuss their results with the Principal Investigator. PI will discuss further with the team if there is any discrepancy.

## Data Availability

Not applicable.

## Acknowledgements

This project is supported by the Florida Department of Health under the contract CODUS.

## Notes

### Competing Interest Statement

The authors have declared no competing interest.

### Author Declarations

For the scoping review we will only use data from published manuscript. The search terms has been included to facilitate replication of the work.

